# Trends and key disparities of obesity among US adolescents: the NHANES from 2007 to 2020

**DOI:** 10.1101/2023.08.06.23293721

**Authors:** Yangyang Deng, Sami Yli-Piipari, Omar El-Shahawy, Kosuke Tamura

**Author notes:** Yangyang Deng and Kosuke Tamura were responsible for the design and data collection, and statistical analysis, the literature review, data interpretation, and manuscript preparation. Sami Yli-Piipari edited the manuscript and provide expertise on the manuscript. Omar El-Shahawy edited the manuscript and helped with interpretation of the results. There are no conflicts of interest to disclose or financial disclosures by any authors.

## Abstract

This study aimed to estimate the trends in the body mass index (BMI) and prevalence of obesity among United States (U.S.) adolescents (10-19 years), and to examine the associations between sociodemographic factors and both BMI and obesity prevalence. The 2007-2020 National Health and Nutrition Examination Survey (NHANES), a nationally representative repeated cross-sectional survey data (n = 9,826) were used. Outcomes included: 1) Mean BMI and 2) obesity (yes/no; defined as BMI ≥ 95% percentile). Sociodemographic variables included age, sex, race/ethnicity, and poverty income ratio (PIR; low-income <1.3, Middle-income ≥1.3 and <3.5, high-income ≥3.5). By accounting for the complex survey design, weighted generalized linear/Poisson models were used to conduct the analyses. Girls constituted 49 % of the sample. From 2007-2008 to 2017-2020, BMI and obesity prevalence increased across various subgroups, including Black and Hispanic adolescents, boys, and those from low- and middle-income families. Girls are a 12% lower likelihood of being obese than boys. Compared to White adolescents, Black and Hispanic adolescents had 22% and 19% greater risk of being obese. Compared to those from high-income families, adolescents from low- and middle-income families had 62% and 47% greater risk of being obese, respectively. The results indicated persistent disparities in obesity prevalence among different race/ethnic and sociodemographic groups. Future obesity intervention should address key disparities by targeting specific race/ethnic adolescents from low-income families and promoting health equality.

## Introduction

The increasing prevalence of childhood obesity in the United States (U.S.) has become a significant public health concern, elevating the risk of developing cardiometabolic comorbidities among U.S. adolescents.(1–4) Recent data from the Centers for Disease Control and Prevention (CDC) (5) showed that the prevalence of obesity among children and adolescents aged 2–19 years increased from 12 million (16.9%) in 2010 to 14.7 million (19.7%) in 2020. These concerning trends have long-term implications for the health and well-being of adolescents as they transition into adulthood.(6–8) The economic burden of obesity in the U.S. healthcare system was estimated to be $173 billion in 2021.(9) Thus, monitoring national trends and examining the association between sociodemographic factors and weight-related outcomes among adolescents are critical to increase awareness and prevent childhood obesity.

Obesity among adolescents (aged 10-19 years) is defined as age- and sex-specific body mass index (BMI) greater than or equal to the 95th percentile based on the 2000 CDC growth chart.(4) Sociodemographic factors, such as age, sex, race/ethnicity, and socioeconomic status, have been associated with the prevalence of childhood obesity.(1, 10–12) For example, a recent study showed that there was a 4.5% increase in obesity prevalence among adolescents aged 12- 19, from 17.7% in 2009-2010to 21.5% in 2017-2020.(13) Research showed a higher prevalence of obesity in boys than girls.(14) This increased rates is particularly pronounced among certain racial and ethnic minorities.(15, 16) The most recent 2020 CDC data indicated that the prevalence of obesity among Black and Hispanic children were 24.8% and 26.2%, respectively, compared to 16.6% among White children.(5) In addition, youth from lower socioeconomic backgrounds are more likely to be obese than those from higher socioeconomic backgrounds,(15, 17) families with lower incomes were 1.39 times more likely to have children who are obese than those from families with higher incomes.(18)

Nevertheless, most studies on sociodemographic factors in relation to childhood obesity were cross-sectional reporting point prevalence estimates,(19–21) with limited studies examining the key disparities in sociodemographic factors with long-term trends in mean BMI and obesity prevalence. Thus, there is highly limited studies on the most recent trends in obesity prevalence among U.S. adolescents aged 10-19 years. To address these gaps, we used a nationally representative sample from the National Health and Nutrition Examination Survey (NHANES) from 2007 to 2020. The aims of this study were two-fold: 1) to illustrate trends in mean BMI and obesity prevalence among U.S. adolescents, and 2) to examine how sociodemographic factors were associated with the mean BMI and obesity prevalence among this sample of U.S. adolescents.

## Methods

### Study sample

This study utilized data from the NHANES from 2007 to 2020,(22) including the cross-sectional survey waves for years 2007-2008, 2009-2010, 2011-2012, 2013-2014, 2015-2016, and 2017-2020. To maintain a nationally representative estimates, the 2017-2020 period included the data until the beginning of the COVID-19 pandemic in March 2020. (5) NHANES uses a stratified multistage sampling method to represent the US population. The sampling frame included a total of 11,357 participants aged 10-19 years. Pregnant participants or those without outcome measurements were excluded from the analysis (n = 1531 [13.5%]), resulting in the final analytical sample of N=9,826 across the six NHANES waves from 2007 to 2020 (eFig. 1 in the Supplement). The NHANES protocol was approved by the National Center for Health Statistics research ethics review board, and all participants provided written informed consent.

**Fig 1.**
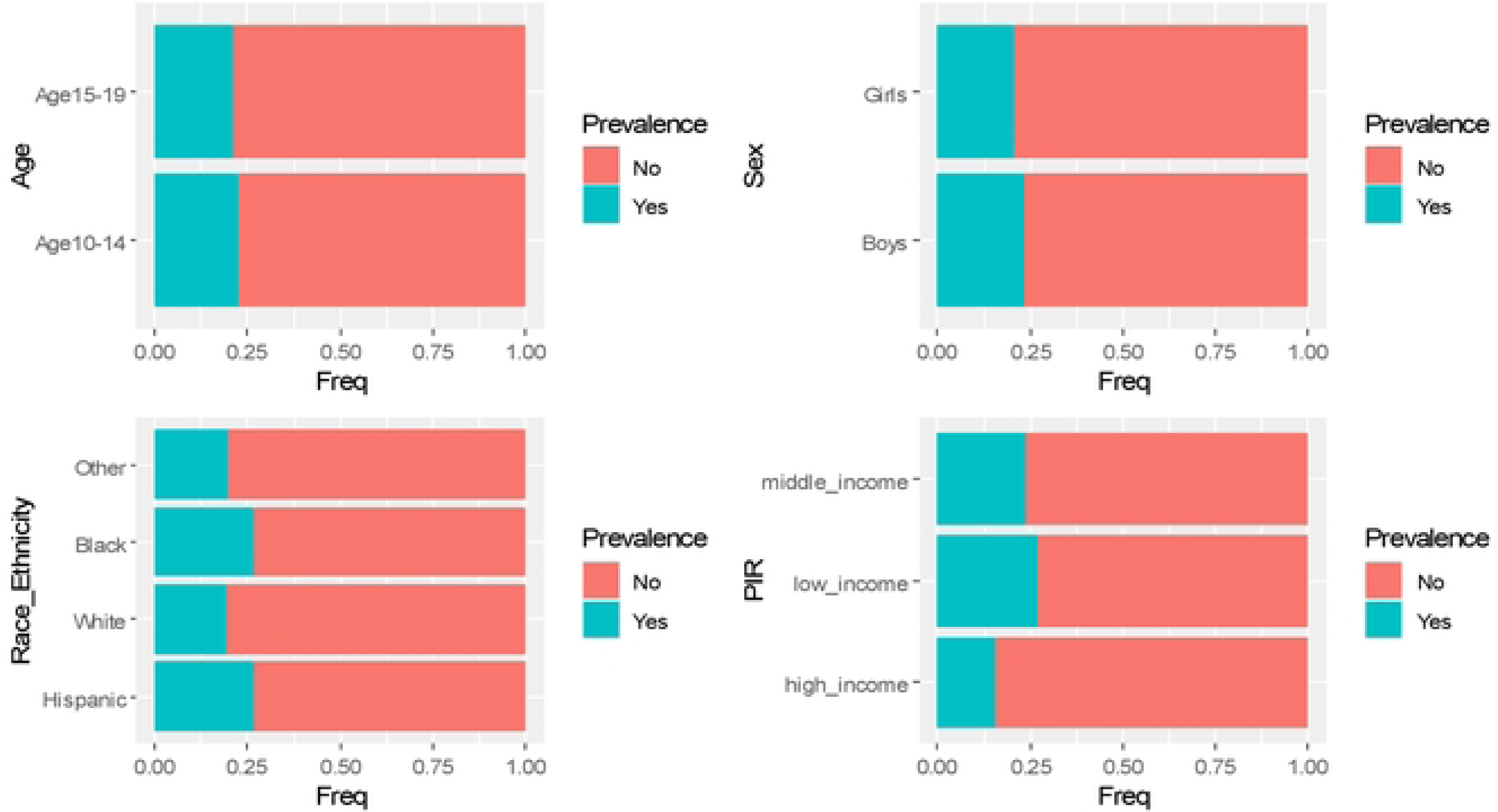
Weighted proportions in obesity prevalence stratify by sex, age and race/ethnicity and poverty income ratio from 2007-2008 to 2017-2020

### Measures

All body measurements were assessed in accordance with a standardized protocol using calibrated instruments.(5) Standing height of the participants was measured using a portal stadiometer and weight was measured using a portal digital weight scale. BMI was calculated by dividing weight in kilograms by squared height in meters. We further calculated their BMI percentile based on the relevant CDC growth charts for children and adolescents, which included ages 10 to 19 years.(23) The adolescent’s obesity were evaluated as their BMI percentile to classify into underweight (less than 5th percentile), normal weight (5th to 84th percentile), overweight (85th to 94th percentile), and obesity (at or above 95th percentile). A binary adolescent’s obesity outcome (Yes, No) was defined as 95th percentile (referent) <BMI ≥ 95th percentile.(23)

Sociodemographic variables included age, sex (girls/boys), race/ethnicity, and poverty income ratio (PIR). Age was self-reported during the interview and was divided into two age groups: 10 to 14 years and 15 to 19 years.(24) Race/ethnicity was categorized as non-Hispanic White (referent), non-Hispanic Black, Hispanic (including Mexican American and other Hispanic), and other (including multi-racial).(25, 26) The PIR is a measure of family income relative to poverty guidelines, which was determined by dividing family income by the poverty threshold specific to a family size and geographic location income levels were defined based on the PIR and coded as low income (PIR <1.3), middle income (PIR ≥1.3 and <3.5), and high income (PIR ≥3.5).(27)

### Statistical analyses

Preparing the data involved several steps, such as removing missing values, examining the distribution, and identifying and removing outliers. Survey analysis procedures were used to derive nationally representative estimates, accounting for sampling weights, strata, and primary sampling units (PSU) in the NHANES complex sampling design.(28) A survey-weighted linear regression model was used to evaluate BMI trends, both overall and stratified by the subgroups (age, sex, race/ethnicity, and PIR), 2007 through 2020.

Subsequently, a survey-weighted Poisson regression model was used to evaluate the prevalence of obesity trends, both overall and by the subgroups (age, sex, race/ethnicity, and PIR), 2007 through 2020. To evaluate the statistical heterogeneity of trends by subgroups, we used a survey-weighted Wald test for an interaction term between survey cycle and sociodemographic factors such as age, sex, race/ethnicity, and PIR. For each survey cycle, we report 95% confidence intervals (CI) for all estimates.

We used multivariate survey-weighted generalized linear/Poisson regression models with robust standard errors to examine the association between sociodemographic factors and mean BMI and obesity prevalence, adjusting for survey wave. Statistical significance was determined by p-value < 0.05 (2-tailed). All analyses were conducted using the R statistical software “Survey” package (e.g., “svymean” and “svyglm”), taking into account the complex sampling design (version 4.2.2; www.r-project.org).

## Results

### Characteristics of study population

The nationally representative sample characteristics from 2007-2008 through 2017-2020, and the characteristics were presented in Table 1. The total sample comprised 9,826 adolescents with mean age =14.29 (SD = ± 2.78), of which 49.0% were girls. The mean BMI increased from 22.85 (95% CI: 22.40-23.29) in 2007-2008 to 23.66 (95% CI: 23.15-24.03) in 2017-2020.

**Table 1.**
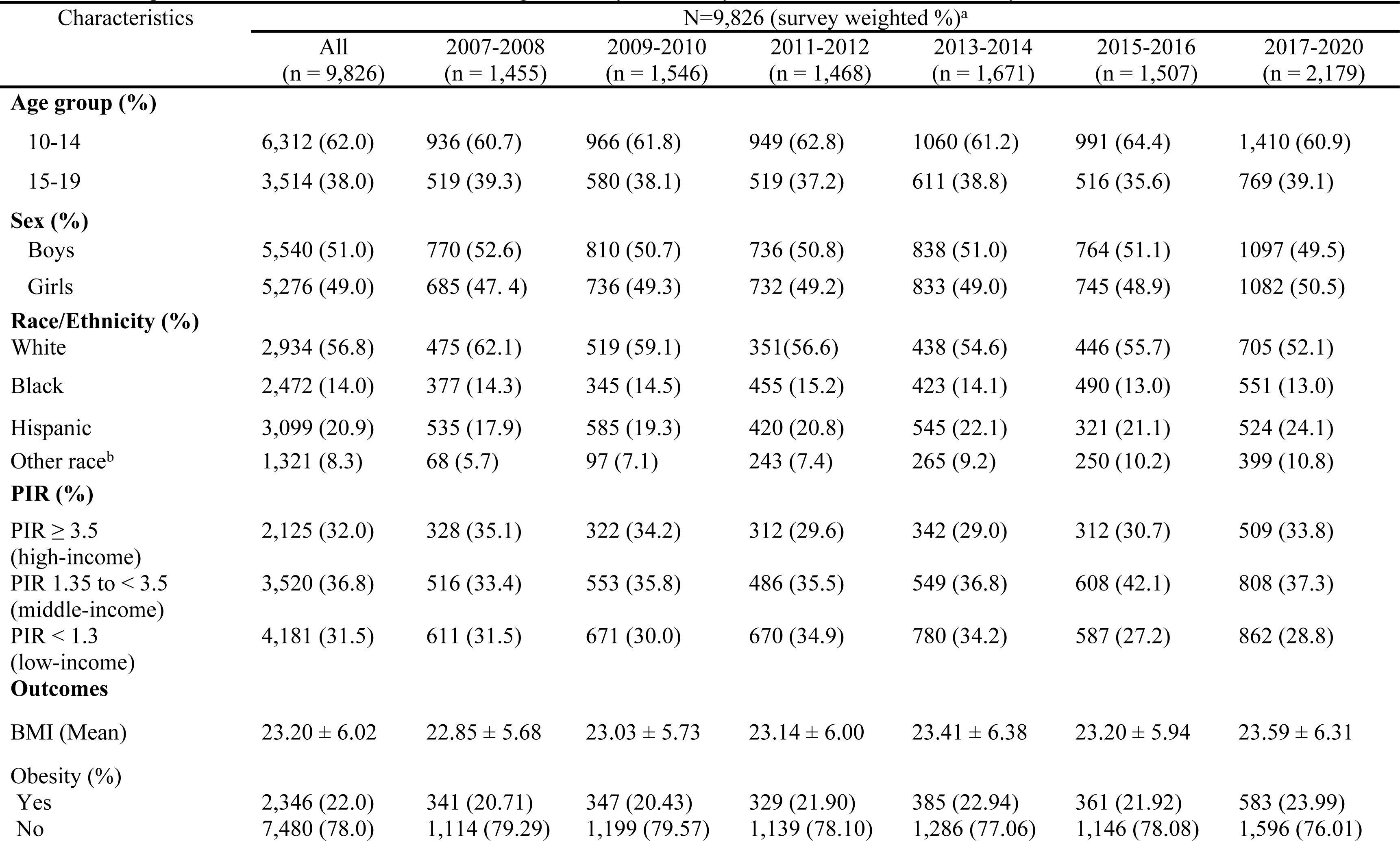
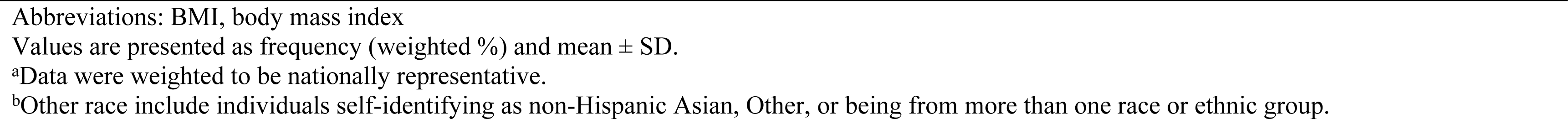
Descriptive table of children and adolescents aged 10-19 years old, by overall, sex, and race/ethnicity from 2007 to 2020.

| Characteristics | N=9,826 (survey weighted %)<sup>a</sup> |  |  |  |  |  |  |
| --- | --- | --- | --- | --- | --- | --- | --- |
|  | All<br>(n = 9,826) | 2007-2008<br>(n = 1,455) | 2009-2010<br>(n = 1,546) | 2011-2012<br>(n = 1,468) | 2013-2014<br>(n = 1,671) | 2015-2016<br>(n = 1,507) | 2017-2020<br>(n = 2,179) |
| **Age group (%)** |  |  |  |  |  |  |  |
| 10-14 | 6,312 (62.0) | 936 (60.7) | 966 (61.8) | 949 (62.8) | 1060 (61.2) | 991 (64.4) | 1,410 (60.9) |
| 15-19 | 3,514 (38.0) | 519 (39.3) | 580 (38.1) | 519 (37.2) | 611 (38.8) | 516 (35.6) | 769 (39.1) |
| **Sex (%)** |  |  |  |  |  |  |  |
| Boys | 5,540 (51.0) | 770 (52.6) | 810 (50.7) | 736 (50.8) | 838 (51.0) | 764 (51.1) | 1097 (49.5) |
| Girls | 5,276 (49.0) | 685 (47.4) | 736 (49.3) | 732 (49.2) | 833 (49.0) | 745 (48.9) | 1082 (50.5) |
| **Race/Ethnicity (%)** |  |  |  |  |  |  |  |
| White | 2,934 (56.8) | 475 (62.1) | 519 (59.1) | 351 (56.6) | 438 (54.6) | 446 (55.7) | 705 (52.1) |
| Black | 2,472 (14.0) | 377 (14.3) | 345 (14.5) | 455 (15.2) | 423 (14.1) | 490 (13.0) | 551 (13.0) |
| Hispanic | 3,099 (20.9) | 535 (17.9) | 585 (19.3) | 420 (20.8) | 545 (22.1) | 321 (21.1) | 524 (24.1) |
| Other race<sup>b</sup> | 1,321 (8.3) | 68 (5.7) | 97 (7.1) | 243 (7.4) | 265 (9.2) | 250 (10.2) | 399 (10.8) |
| **PIR (%)** |  |  |  |  |  |  |  |
| PIR ≥ 3.5<br>(high-income) | 2,125 (32.0) | 328 (35.1) | 322 (34.2) | 312 (29.6) | 342 (29.0) | 312 (30.7) | 509 (33.8) |
| PIR 1.35 to < 3.5<br>(middle-income) | 3,520 (36.8) | 516 (33.4) | 553 (35.8) | 486 (35.5) | 549 (36.8) | 608 (42.1) | 808 (37.3) |
| PIR < 1.3<br>(low-income) | 4,181 (31.5) | 611 (31.5) | 671 (30.0) | 670 (34.9) | 780 (34.2) | 587 (27.2) | 862 (28.8) |
| **Outcomes** |  |  |  |  |  |  |  |
| BMI (Mean) | 23.20 ± 6.02 | 22.85 ± 5.68 | 23.03 ± 5.73 | 23.14 ± 6.00 | 23.41 ± 6.38 | 23.20 ± 5.94 | 23.59 ± 6.31 |
| **Obesity (%)** |  |  |  |  |  |  |  |
| Yes | 2,346 (22.0) | 341 (20.71) | 347 (20.43) | 329 (21.90) | 385 (22.94) | 361 (21.92) | 583 (23.99) |
| No | 7,480 (78.0) | 1,114 (79.29) | 1,199 (79.57) | 1,139 (78.10) | 1,286 (77.06) | 1,146 (78.08) | 1,596 (76.01) |

Abbreviations: BMI, body mass index
Values are presented as frequency (weighted %) and mean ± SD.
<sup>a</sup>Data were weighted to be nationally representative.
<sup>b</sup>Other race include individuals self-identifying as non-Hispanic Asian, Other, or being from more than one race or ethnic group.

Furthermore, the prevalence of obesity increased from 20.71% (95% CI: 17.16-24.27) in 2007-2008 to 23.99% (95% CI: 21.31-26.67) in 2017-2020. In the overall analyses, there were no significant trends in the mean BMI changes and the overall prevalence of obesity (Table 1, eTables 1 and 2). Figure 1 showed overall obesity prevalence weighted proportions, stratified by age, sex, race/ethnicity, and PIR. Notably, Black and Hispanic adolescents showed higher obesity prevalence proportion rates than other race/ethnicity. Moreover, adolescents from low-income demonstrated the highest prevalence.

The Wald test analysis did not reveal any significant interaction between survey cycle and both BMI and obesity prevalence concerning age groups and sex. However, a significant interaction effect between survey cycle and both BMI and obesity prevalence was observed across various race/ethnic and PIR groups (eTables 1 and 2).

### Sociodemographic stratified trends and disparities in the BMI and prevalence of obesity

For 15-19 age group (with 10-14 as the reference group), mean BMI level significantly increased from 24.48 (95% CI: 23.84-25.13) in 2007-2008 to 25.53 (95% CI: 24.94-26.12) in 2020 (p<0.001) (efig2 (a), and eTable 1 in the Supplement). However, the prevalence of obesity did not significantly increase from 20.62 (95% CI: 16.98-24.25) to 22.11 (95% CI: 18.29-25.94) (eTable 2 in the Supplement). For girls (boys [reference]), mean BMI levels did not significantly increase from 22.11 (95% CI: 22.33-23.50) in 2007-2008 to 23.78 (95% CI: 23.23-24.33) in 2017-2020 (p>0.05) (efig2 (b), and eTable 1 in the Supplement). However, there is a significant the prevalence of obesity increases for girls (boys [reference]), from 18.24 (16.08-20.41) in 2007-2008 to 22.11 (95% CI: 18.47-26.22) in 2017-2020 (p<0.01).

Among race/ethnic groups, Black adolescents had the highest increase in mean BMI from 23.90 (95% CI: 23.28-24.53) to 25.92 (95% CI: 24.28-25.56) (p<0.001) between 2007 and 2020 (efig 2 [c], in the Supplement). Hispanic adolescents also had the increase in mean BMI from 22.54 (95% CI: 22.89-24.18) to 24.07 (95% CI: 23.31-24.83) (p<0.001). Yet, there was no significant trend in the Other race group. Similarly, for the obesity prevalence (fig 2 [c]), Black adolescents also had the highest increase, with a rise of 5.7% from 24.96 (95% CI: 22.07-33.45) to 30.62 (95% CI: 27.18-34.06) (p<0.001) in 2007-2020. In addition, Hispanic adolescents had an increase from 24.21 (95% CI: 20.78-28.67) to 29.05 (95% CI: 22.97-35.13) (p<0.001).

**Fig 2.**
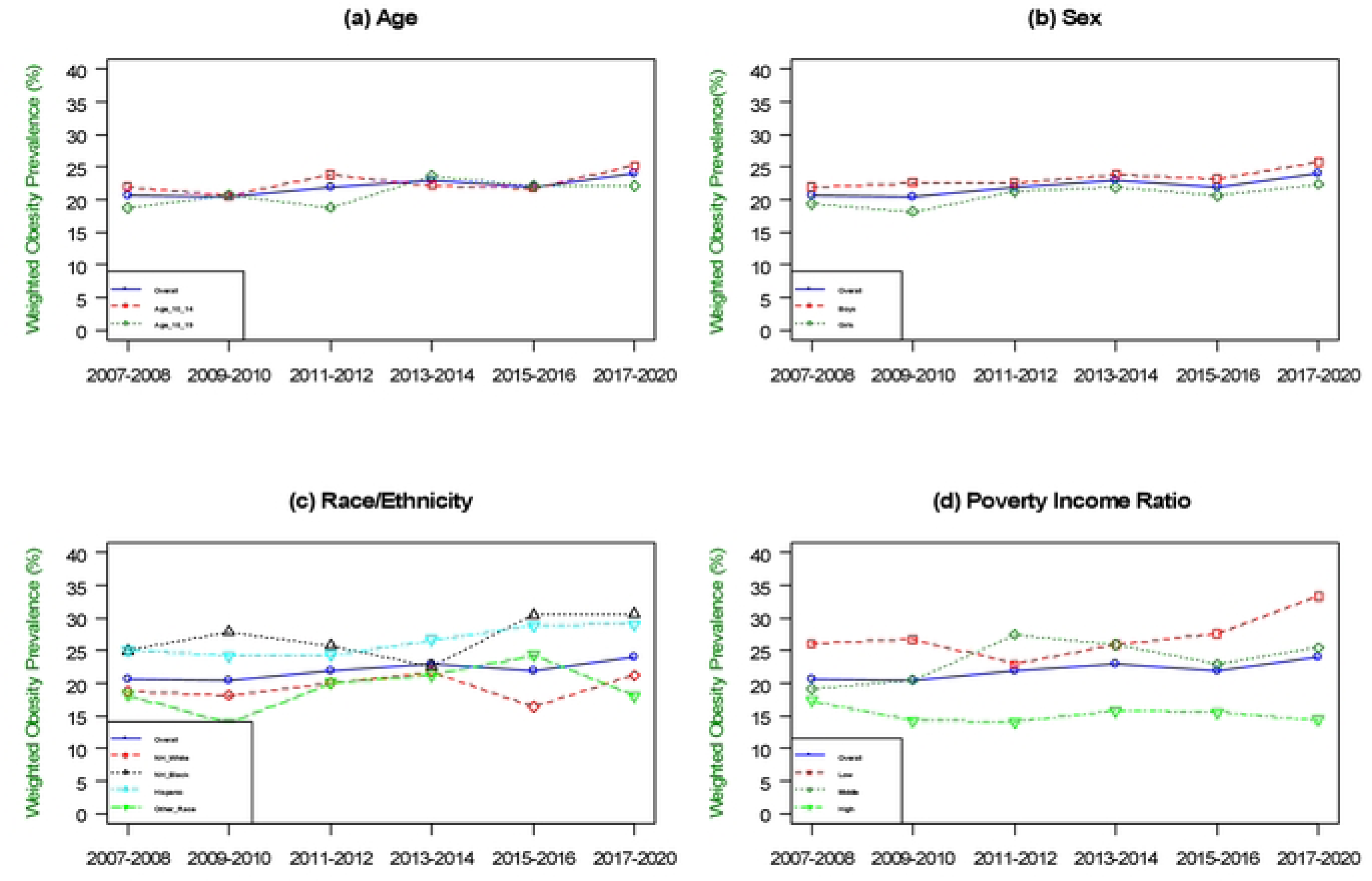
Trends in prevalence of obesity among US adults (aged 10-19), NHANES 2007-2020 by (a)Age, (b) Sex, (c) Race/ethnicity, and (d) Poverty Income Ratio.

However, there was no significant trend in the Other race.

Comparing adolescents from high income families (PIR ≥ 3.5), adolescents from low income (PIR < 1.3) and middle-income families (PIR 1.3 to < 3.5) had significant increases in mean BMI (efig 2 [d], in the Supplement). Specifically, the low-income group had a mean BMI of 23.57 (95% CI: 22.83-24.31), and the middle-income group had a mean BMI of 22.84 (95% CI: 22.10-23.59) in 2007-2008 to 24.82 (95% CI: 24.22-25.42), and 23.67 (95% CI: 22.96-24.38) in 2017-2020. The prevalence of obesity also increased significantly in the middle-income group, from 20.52 (95% CI: 16.91-24.14) in 2007-2008 to 25.45 (95% CI: 21.32-29.58) in 2017-2020.

However, there was a significant decrease in the prevalence of obesity in the high-income group with PIR ≥ 3.5, from 17.31 (95% CI: 17.31-22.21) in 2007-2008 to 14.44 (95% CI: 11.50-17.38) in 2017-2020.

### Association of sociodemographic factors with BMI and obesity

Table 2 showed that age, sex, race/ethnicity, and PIR were significantly associated with mean BMI. Adolescents aged 15-19 years had a higher BMI (β=3.27, 95% CI: 2.91-3.62) compared to ones aged 10-14 years. Black and Hispanic adolescents had a higher BMI (β=1.10, 95% CI: 0.59-1.62, and β=0.69, 95% CI: 0.29-1.09, respectively), compared to White counterparts. Furthermore, low and middle-income adolescents had a higher BMI (β=1.43, 95% CI: 1.04-1.83, β=1.09, 95% CI: 0.60-1.60, respectively) than ones from high-income families.

**Table 2.** Weighted generalized linear/Poisson models s for obesity among US children and dolescents aged 10-19 years old, 2007-2020.

|  | Mean BMI |  | Prevalence of obesity |  |
| --- | --- | --- | --- | --- |
| | Adjusted $\beta$<br>(95%CI) | P-value | Adjusted prevalence<br>ratio (95% CI) | P-value |
| **Age group** |  |  |  |  |
| 10-14 | reference |  | 1.00 (reference) |  |
| 15-19 | 3.27 (2.91, 3.62) | <0.001 | 0.95 (0.85, 1.05) | 0.166 |
| **Sex** |  |  |  |  |
| Boys | reference |  | 1.00 (reference) |  |
| Girls | 0.23 (-0.08, 0.53) | >0.05 | 0.88 (0.81, 0.96) | <0.01 |
| **Race/Ethnicity** |  |  |  |  |
| Non-Hispanic white | Reference |  | 1.00 (reference) |  |
| Non-Hispanic black | 1.10 (0.59, 1.62) | <0.001 | 1.22 (1.06, 1.40) | <0.01 |
| Hispanic | 0.69 (0.29, 1.09) | <0.001 | 1.19 (1.05, 1.36) | <0.01 |
| Other Race<sup>a</sup> | -0.37 (-0.95, 0.22) | >0.05 | 0.93 (0.82, 1.04) | >0.05 |
| **Poverty Income Ratio** |  |  |  |  |
| PIR ≤ 3.5<br>(high-income) | reference | | 1.00 (reference) | |
| PIR < 1.3<br>(low-income) | 1.43 (1.04, 1.83) | <0.001 | 1.62 (1.39, 1.90) | <0.001 |
| PIR 1.3 to <3.5<br>(middle-income) | 1.09 (0.60, 1.60) | <0.001 | 1.47 (1.24, 1.76) | <0.001 |

Models were adjusted for survey year.
<sup>a</sup>Other race include individuals self-identifying as non-Hispanic Asian, Other, or being from more than one race or ethnic group.

Table 2 showed there was no statistically significant relationship between age and obesity prevalence. Girls had a lower likelihood of obesity compared to boys (AOR=0.88; 95% CI, 0.81- 0.96, p<0.01). Furthermore, Black and Hispanic adolescents had a higher risk of obesity than White adolescents (Adjusted odds ratio Black; (AOR)=1.22; 95% CI: 1.06-1.40, p<0.01; and Hispanic; AOR=1.19; 95% CI, 1.05-1.36, p<0.01, respectively). Additionally, adolescents with a low and middle income were 62% and 47% more likely to be obese, compared to ones from high income groups.

## Discussion

We examined trends in BMI and obesity prevalence among US adolescents using nationally representative NHANES data from 2007-2008 to 2017-2020. This study provided the most up-to-date available US nationally representative trends over the decade. Our findings indicated that adolescent obesity continued to increase in the US over the past decade. We found the increase in both mean BMI and obesity prevalence from 2007-2008 to 2017-2020 among several subgroups of adolescents, including boys and girls, Black and Hispanic adolescents (compared to White counterparts), and those from low and middle-income backgrounds (compared to high-income background). In contrast, we found a decrease in obesity prevalence among those from high-income groups. Our analysis also revealed persistent disparities among certain racial minorities (Black and Hispanic adolescents) and those in low and middle-income groups (PIR ≤ 1.3 and PIR 1.35 to < 3.5) who had greater odds of being obesity. These findings underscore the importance of targeted interventions to address the growing obesity epidemic among US adolescents, particularly those from minority groups and lower socioeconomic status backgrounds.

The BMI increased from 22.85 kg/m^2^ in 2007-2008 to 23.59 kg/m^2^ in 2017-2020 among adolescents aged 10-19, while the prevalence of obesity increased by 3.7% from 20.7% to 24% over the same time period. Our findings were consistent with prior research indicating that childhood obesity remained high in the U.S., with approximately 1 in 5 children affected 12- to 19-year-olds.(29) Notably, our study provides updated data on this trend compared to 2017- 2020. In addition, our study findings are consistent with a recent study that reported a 4.5% increase in obesity prevalence among adolescents aged 12-19 from 2009-2010 to 2017-2020, yet presented only unadjusted prevalence estimates,(13) Our study included a longer time span than these previous studies through a robust multivariate survey-weighted data analysis, with adjusting for key sociodemographic factors and survey cycle. Our findings demonstrated disparities in race/ethnic groups and income-levels leading to obesity among U.S. adolescents. Additionally, compared to girls, boys had a higher prevalence of obesity from 22.0% in 2007- 2008 to 25.3% in 2017-2020. This finding was consistent with previous study that the prevalence was slightly higher in boys.(30) Considering sex will enable us to develop more effective and tailored strategies to improve the overall well-being and outcomes of this vulnerable population.

We found that the prevalence of obesity continues to increase among certain race/ethnic groups, as well as low-income groups. Compared to White counterparts, Black and Hispanic adolescents had higher obesity rates in 2017-2020 (30.62% and 29.05%, respectively). These findings are consistent with previous studies,(12, 30, 31) that Black and Hispanic adolescents as having the highest obesity rates among all minority groups. Other races including Asian adolescents had a lower prevalence of obesity in all age and sex groups, which is also in line the with earlier research.(32) However, it is important to note that Asians have different body compositions, and Asian children with a normal BMI range, meant for White children may still be considered overweight or obese according to Asian-specific criteria.(33) In addition, we found a decreasing trend from 17.31% in 2007-2008 to 14.44% in 2017-2020 among high income groups. Conversely, low-income (PIR < 1.3) and middle income (PIR 1.3 to < 3.5) had significant increases in mean BMI and obesity prevalence were observed for both groups.

Consistent with the pervious result showing that adolescents from low-income families had a greater chance of being obese,(15) our study highlights a concerning trend of rising obesity rates among adolescents from low- and middle-income groups.(34) Since adolescents from low-income families often encounter challenges, such as limited access to healthy foods and opportunities for physical activity,(35) they may face obstacles in making healthier choices than those from high-income families.(36, 37) Our overall analyses examining association beween sociodemograhic factors and BMI and obesity revealed persistent disparities among Black and Hispanic adolescents. Black and Hispanic adolescents had 23% and 19% greater odds of being obese compared to White counterparts. Adolescents from low and middle-income groups had higher odds of obesity, while the high-income group had lower odds. These results align with previous studies,(38) indicating that socioeconomic inequalities play a significant role in shaping access to neighborhood and social resources. Furthermore, a previous systematic review demonstrated that approximately 40% of studies have identified a widening socioeconomic inequality gap since 2000.(39) A possible explanation may be that these disparities in obesity prevalence among different race/ethnic and income groups may be linked to various factors such as residing in unsafe neighborhoods that limit opportunities (40) and limited access to healthy food options.(41) Furthermore, another study indicated that adolescents from low socioeconomic neighborhoods are more exposed to poverty, crime, low social cohesion, higher levels of air pollution, lack of green spaces, and poor quality of the built environment, which can increase the likelihood of obesity.(42) Adolescence is a critical period for preventing obesity, the results of our study underscore the substantial disparities in the prevalence of obesity existing among youth.

Therefore, future research should address these disparities and promote health equality.

The strengths of this study included assessment of the most up to-date available data from NHANES to evaluate trends over the past 13 years. In addation, while the survey data is mostly self-reported and are subject to misclassification, a strength of the our approach is that the weight and height data collected from NHANES were measured by professional data collectors rather than self-report, increasing the accuracy and reliability of the results by reducing personal bias.(17) Another strength is the large sample size, which provided sufficient statistical power to examine the race- and sex-specific mean BMI and prevalence of obesity, despite not accounting for Asian American adolecents. Lastly, this study employed a rigorous multivariate survey-weighted data analysis, and accounting for key sociodemographic factors and survey cycles. This methodological approach significantly enhances the depth and reliability of the findings, strengthening the validity of the study’s conclusions.

There are several limitations. First, the NHANES data are based on repeated cross-sectional surveys, which means that we cannot examine within-child changes over time or imply causality. However, this approach still provides a valuable snapshot of obesity prevalence among US adolescents over time. In addition, including Asian American adolescents in the other group in our analysis raises questions about the validity of current reference ranges for defining obesity.(32)

## Conclusions

Despite efforts in public health policies and increasing awareness, the prevalence of obesity among children remains high and continues to increase among Black and Hispanic adolescents. Future research should focus on addressing key disparities suggested through this analysis and promoting health equality. The findings of our study suggest that future obesity interventions should be target to specific race/ethnic minorities and low-income families to prevent obesity. This could involve developing policies and interventions addressing social determinants of health, such as improving access to healthy food options, creating safe environments for physical activity, and promoting culturally appropriate community-based initiatives that support healthy lifestyles.(42)

## Data Availability

All relevant data are within the manuscript and its Supporting Information files.

## Acknowledgements

Funding for the Socio-Spatial Determinants of Health (SSDH) Laboratory is provided through the Division of Intramural Research at the National Institute on Minority Health and Health Disparities (NIMHD), the National Institute of Health (NIH), and the NIH Distinguished Scholars Programs. Yangyang Deng is supported by the NIH Postdoctoral Intramural Research Training Award.

The views of this study are those of the authors and do not necessarily represent the views of the NIMHD, the NIH, and the U.S. Department of Health and Human Services.

No financial disclosures were reported by the authors of this paper.

## Supporting information

S Table 1. BMI Changes in Adolescents aged 10-19 years old, stratify by overall, sex, age, race/ethnicity, and poverty ratio from 2007 to 2020 (n=9,816)

S Table 2. Prevalence of Obesity in Adolescents aged 10-19 years old, stratify by overall, sex, age, race/ethnicity, and poverty ratio from 2007 to 2020 (n=2,346)

S Fig 1. Sample size flowchart

S Fig 2. Trends in mean BMI obesity among US adults (aged 10-19), NHANES 2007-2020 by Sex, Age, race/ethnicity and Poverty income Ratio (Details see in Supplemental Table 1).

